# High School Sports During the CoVID-19 Pandemic: The Impact of Sport Participation on the Health of Adolescents

**DOI:** 10.1101/2021.02.07.21251314

**Authors:** Timothy A. McGuine, Kevin Biese, Scott J. Hetzel, Allison Schwarz, Claudia L. Reardon, David R. Bell, M. Alison Brooks, Andrew M. Watson

## Abstract

**Context:** During the fall of 2020, some high schools across the US allowed their students to participate in interscholastic sports while others cancelled or postponed their sport programs due to concerns regarding CoVID19 transmission. It is unknown what effect this has had on the physical and mental health of student athletes.

**Objective:** Identify the impact of playing a sport during the CoVID19 pandemic on the health of student athletes.

**Design:** Cross-sectional study.

**Setting:** Sample recruited via email.

**Patients or Other Participants:** 559 Wisconsin high school athletes (age=15.7±1.2 yrs., female=44%) from 44 high schools completed an online survey in October 2020. A total of 171 (31%) athletes played (PLY) a fall sport, while 388 (69%) did not play (DNP).

**Main Outcome Measure(s):** Demographics included: sex, grade and sports played. Assessments included the General Anxiety Disorder-7 Item (GAD-7) for anxiety, Patient Health Questionnaire-9 Item (PHQ-9) for depression, the Pediatric Functional Activity Brief Scale (PFABS) for physical activity, and the Pediatric Quality of Life Inventory 4.0 (PedsQL) for quality of life. Univariable comparisons between the two groups were made via t-tests or chi-square tests. Means for each continuous outcome measure were compared between the groups by ANOVA models that controlled for Age, Sex, Teaching method (Virtual, Hybrid, or In-person), and the % of students eligible for free lunch.

**RESULTS:** PLY group participants were less likely to report moderate to severe symptoms of anxiety (PLY=6.6%, DNP=44.1%, p<0.001) and depression (PLY=18.2%, DNP=40.4%, p<0.001). PLY athletes reported higher (better) PFABS scores (mean: [95%CI]), (PLY=23.2[22.0,24.5], DNP=16.4[15.0,17.8], p <0.001) and higher (better) PedsQL total scores (PLY=88.4[85.9,90.9], DNP=79.6[76.8,82.4], p <0.001).

**CONCLUSIONS:** Adolescent athletes who played a sport during the CoVID19 pandemic reported fewer symptoms of anxiety and depression, as well as higher physical activity and quality of life scores compared to adolescent athletes who did not play a sport.

**Key points:**

1. High school students who played a sport during the CoVID-19 pandemic in the fall of 2020 were less likely to report anxiety and depression symptoms than athletes who did not play a sport.
2. High school students who played a sport during the CoVID-19 pandemic in the fall of 2020 reported higher physical activity and quality of life scores compared to high school athletes who did not play a sport.
3. Participation in high school sports may have significant physical and mental health benefits for US adolescent athletes during the CoVID-19 pandemic.

## INTRODUCTION

An estimated 8.4 million US high school students participate in interscholastic athletics each year.^1^ Adolescent sport participation is recognized to have profound positive influences on the health and well-being of adolescent students as evidenced by higher academic achievement, greater levels of physical activity and decreased levels of anxiety and depression compared to students who do not participate in athletics.^2-6^ Additionally, research has shown that high school sport participation is one of the most important factors for life-long physical activity and health.^7-12^

During the spring of 2020, CoVID-19, the disease caused by the novel SARS-CoV-2 coronavirus, reached pandemic levels in the US. Schools were closed and high school sports were cancelled to slow the spread of the disease. Experts have suggested that while necessary to slow the community spread of the virus, the CoVID-19 mitigation strategies may nonetheless have profound mental and physical health consequences for students.^13-18^ For example, experts have indicated that the CoVID-19 pandemic will make it more likely that youth will engage in sedentary activities and increase the prevalence of childhood obesity.^19-23^

Research has also shown that females, older athletes, team sport participants and athletes from areas with higher levels of poverty reported more symptoms of anxiety and depression, as well as lower levels of physical activity and health related quality of life (HRQoL) in May 2020 during the widespread shutdown.^24^ Another study reported that adolescent athletes in May 2020 reported more mental health symptoms, as well as lower physical activity and HRQoL scores compared to similar samples of adolescent athletes prior to the CoVID-19 pandemic. ^25^

During summer 2020, school districts across the US made determinations regarding whether to allow interscholastic sports to resume based on input from various health and sport associations while recognizing that various sports would possibly pose varying risks for the transmission of CoVID-19 depending on the nature of the sport. ^26-28^ During fall 2020, fourteen states in the US (27%) allowed full fall sport participation, and 30 states (60%) allowed modified sport participation, with the remaining 6 states and the District of Columbia (13%) not allowing any interscholastic athletic participation.^29^ In addition, within each state that allowed full or modified participation, individual school districts were allowed to determine which sports they offered. Some schools allowed all their fall sports, while other districts sponsored a portion of their sports while postponing or cancelling other sports.^29^

A drawback to the recent studies regarding the impact of CoVID-19 on the health of adolescent athletes is the difficulty discerning if the health changes reported were primarily due to restrictions on sport participation or due to other factors such as the lack of in-person school attendance, the increased economic uncertainty or concerns about contracting the SARS-CoV-2 coronavirus. There is a need, therefore, to determine if sport participation independently affects the health of adolescent athletes during the CoVID-19 pandemic.

We are not aware of research to date that has specifically documented how sport participation during the CoVID-19 pandemic affects the mental and physical health of adolescent athletes. This information may assist sports medicine providers, school administrators and health care policy experts with implementing strategies to improve the short-term and long-term mental and physical health of adolescent athletes in the months and years to come as we transition from the CoVID-19 pandemic.^30,31^Therefore, the purpose of this study was to measure the impact of sport participation on the mental and physical health of adolescent athletes during the CoVID-19 pandemic. To measure this impact, we compared self-report data on anxiety, depression, physical activity and HRQoL for a cohort of athletes who played a sport with a similar cohort of athletes who did not play a sport in Wisconsin during fall 2020. We hypothesize that adolescent student athletes who played a sport will report significantly better mental health, physical activity and HRQoL than athletes who did not play a sport.

## METHODS

This study was approved by the University of Wisconsin Health Sciences Institutional Review Board in September 2020.

Wisconsin allowed individual school districts to determine whether to sponsor interscholastic teams during the fall of 2020.^32^ The traditional fall sports (prior to the CoVID-19 pandemic) offered in Wisconsin high schools included cross country, football, volleyball (boys and girls), golf (girls), swim (boys and girls), tennis (girls), and soccer (boys). Approximately 305 (60%) of the 510 schools opted to sponsor teams for all fall sports, while 104 (20%) opted to offer a limited number of sports, and the remaining 101 (20%) offered no fall sports at their schools.

The sports offered most often by schools included boys’ and girls’ cross country (78%) and girls’ golf (72%). The sports offered least often included football (54%), boys’ soccer (50%) and girls’ swimming (50%).^33^

Wisconsin high school athletes (male and female, grade: 9–12, age: 13-19) were recruited to participate in the study by completing an anonymous online survey in October 2020. Emails were sent to athletic trainers and coaches from 44 schools to solicit their athletes to participate in the study. The survey included 69 items and included a section to solicit demographic information, followed by 3 validated instruments used to measure physical activity, mental health and HRQoL in adolescents. Demographic responses regarding the participant’s age, grade and school name, as well as any high school sport in which they competed during the fall and planned to compete in if the sport was offered by the school during the winter and spring of the 2020/21 school year were collected. The remainder of the survey consisted of an assessment of mental health, physical activity level and HRQoL.

### Mental health

The General Anxiety Disorder-7 Item (GAD-7) and Patient Health Questionnaire-9 Item (PHQ-9) surveys were used to evaluate anxiety and depression symptoms, respectively.^34^ The questionnaires ask participants to rate the frequency of anxiety or depression symptoms experienced in the past two weeks. The GAD-7 scale is a valid, reliable and sensitive measure of anxiety symptoms and is able to differentiate between mild and moderate GAD in adolescents.^35^ Scores range from 0-21 with a higher score indicating increased anxiety. In addition to the total score, GAD-7 categorical scores of 0– 4, 5-9, 10–14, and 15–21 correspond to no, mild, moderate, and severe anxiety symptoms, respectively.^36^ The PHQ-9 is a 9-item screening questionnaire for depression symptoms with scores ranging from 0-27 with a higher score indicating a greater level of depression. The PHQ-9 has demonstrated high sensitivity and specificity for depression screening in adolescent patients aged 13 to 17 years.^37^ In addition to the total score, PHQ-9 categorical scores of 0–4, 5–9, 10–14, 15–19 and ≥ 20 correspond to minimal or none, mild, moderate, moderately severe and severe depression symptoms, respectively.^38^

### Physical Activity

Physical activity level was assessed with the Hospital for Special Surgery Pediatric Functional Activity Brief Scale (PFABS). This validated 8-item instrument was designed to measure the activity of active children between 10 and 18 years old for the past month. Scores range from 0 to 30 with a higher score indicating greater physical activity.^39,40^

### Health Related Quality of Life

HRQoL was measured with Pediatric Quality of Life Inventory 4.0 (PedsQL). The 23-item PedsQL questionnaire assesses HRQoL for the previous 7 days. A physical summary score (physical function) and psychosocial (a combination of emotional, social and school function) summary score, as well as the total PedsQL scores can be calculated, with scores ranging from 0 to 100 and a higher score indicating greater HRQoL. The PedsQL has been validated for use in children ages 2 to 18.^41,42^

### Statistical analyses

Statistical analyses were performed for participants who provided a valid, complete survey. Participants were excluded if they did not complete the entire survey, were not in grades 9 -12, or indicated they did not plan to play interscholastic sports at their school. Participants’ demographic variables were summarized (mean [SD] or N [%]) overall and by study participants for sex and fall sport participation. Participants were classified as playing a fall sport (PLY) or as not playing a fall sport (DNP).

The characteristics for the schools attended by the participants included the type of instructional delivery method and the % of students eligible for free or reduced lunch. The type of instructional delivery method (online only, in person or hybrid [a combination of in person and online]) was determined by reviewing information on each school’s website. The % of the students eligible for free or reduced lunch for each school was obtained from the publicly available data available online through the Wisconsin Department of Public Instruction.^43^

Means (mean, [95%CI]) for each continuous outcome measure were reported and compared between fall sport participation groups by ANOVA models that controlled for age, sex, type of teaching method and % of students who were eligible for free or reduced lunch. Ordinal logistic regression models were used to estimate the percentages of level of depression (PHQ-9) and anxiety (GAD-7) by group. These models controlled for the same covariates listed above. All tests had a 0.05 significance level. Analyses were conducted in R for statistical computing version 3.5.

## RESULTS

A total of 559 high school athletes (age = 15.7±1.2 yrs., female = 43.6%, male = 56.4%) completed the survey. Due to the convenience sampling design, information regarding the response rate was unavailable. Three hundred eighty-eight (69.4%) participants reported they did not play (DNP) an interscholastic sport at their school, while 171 (30.6%) reported they did play (PLY) an interscholastic sport. The majority (n = 257, 66.2%) of the participants in the DNP group attended schools that cancelled all fall sports, and 91.4% (n = 355) attended schools that delivered all instruction online. The % of students eligible for free or reduced lunch (mean ± SD) for the schools was 25.9 ± 10.3%. One hundred forty-eight (86.5%) of the participants in the PLY group attended schools that offered all fall sports, with 40.3% (n = 69) attending school in-person. Participants in the PLY group were most likely to report playing volleyball (n = 66, 38.6%), football (n = 53, 31%) and boys’ soccer (n = 22, 12.9%). Participants in the DNP group most commonly reported that they had intended to play football (n = 160, 41.2%), and volleyball (n = 51, 13.1%). Seventy nine participants (20.4%) did not play a fall sport but intendede to play a high school winter or spring sport. A summary of the participant characteristics is found in Table 1.

**Table 1.** Participant Characteristics for Adolescent Athletes Who Did or Did Not Play a High School Sport in the Fall 2020

| Variable | All Participants<br>(N = 559) | Did Not<br>Play a Fall Sport<br>(n = 388) | Did<br>Play a Fall Sport<br>(n = 171) | P-value |
| --- | --- | --- | --- | --- |
| **Age** | 15.7 (1.2) | 15.7 (1.2) | 15.7 (1.2) | 0.895 |
| **Sex** |  |  |  | < 0.001 |
| Female | 244 (43.6%) | 151 (39.0%) | 93 (54.7%) |  |
| Male | 313 (56.3%) | 236 (61.0%) | 77 (45.3%) |  |
| **Grade** |  |  |  | 0.173 |
| 9 | 130 (23.3%) | 90 (23.2%) | 40 (23.4%) |  |
| 10 | 167 (29.9%) | 116 (29.9%) | 51 (29.8%) |  |
| 11 | 145 (25.9%) | 109 (28.1%) | 36 (21.1%) |  |
| 12 | 117 (20.9%) | 73 (18.8%) | 44 (25.7%) |  |
| **Schools** | 44 | 23 | 25 | -- |
| **Instructional delivery method** |  |  |  | < 0.001 |
| Online | 374 (66.9%) | 355 (91.4%) | 19 (11.1%) |  |
| Hybrid (in person and online) | 113 (20.2%) | 30 (7.7%) | 83 (48.5%) |  |
| In person | 72 (12.8) | 3 (0.8%) | 69 (40.3%) |  |
| **% Students eligible for free or reduced lunch** | 25.9 (10.3) | 22.9 (7.0) | 34.0 (13.3) | < 0.001 |
| **Planned Fall Sport Participation** |  |  |  |  |
| Cheer / Dance | 23 (4.1%) | 20 (5.2%) | 3 (1.8%) |  |
| Cross Country | 41 (7.3%) | 25 (6.4%) | 16 (9.4%) |  |
| Football | 213 (38.1%) | 160 (41.2%) | 53 (31.0%) |  |
| Soccer (boys) | 53 (9.5%) | 31 (8.0%) | 22 (12.9%) |  |
| Swim (girls) | 20 (3.6%) | 19 (4.9%) | 1 (0.6%) |  |
| Tennis (girls) | 13 (2.3%) | 3 (0.8%) | 10 (5.8%) |  |
| Volleyball | 117 (20.9%) | 51 (13.1%) | 66 (38.6%) |  |
| None | 79 (14.1%) | 79 (20.4%) | 0 (0.0%) |  |
| **Planned Winter or Spring Sport Participation** |  |  |  |  |
| Baseball | 103 (18.4%) | 72 (18.6%) | 31 (18.1%) |  |
| Ice Hockey | 22 (3.9%) | 18 (4.6%) | 4 (2.3%) |  |
| Lacrosse | 32 (5.7%) | 32 (8.2%) | 0 (0.0%) |  |
| Soccer (girls) | 205 (36.7%) | 123 (31.7%) | 82 (48.0%) |  |
| Softball | 66 (11.8%) | 33 (8.5%) | 33 (19.3%) |  |
| Swim (boys) | 4 (0.7%) | 3 (0.8%) | 1 (0.6%) |  |
| Tennis (boys) | 7 (1.3%) | 7 (1.8%) | 0 (0.0%) |  |
| Track | 120 (21.5%) | 72 (18.6%) | 48 (28.1%) |  |
| Wrestling | 35 (6.3%) | 27 (7.0%) | 8 (4.7%) |  |
| Other sport^ | 24 (4.3%) | 21 (5.4%) | 3 (1.8%) |  |

^ Other sport includes: Bowling, Gymnastics, Rugby, Power lifting, Skiing (alpine and downhill).

### Mental Health

The PLY participants were more likely to report GAD-7 symptom scores of 0 to 4, indicating no or minimal anxiety (PLY = 80.1% vs DNP = 26.4%), whereas the DNP athletes were more likely to report scores of 10 to 21, indicating more moderate to severe anxiety than the PLY group (DNP = 44.1% vs PLY = 6.6%) as shown in Figure 1. The PLY participants were more likely to report PHQ-9 scores 0 to 4, indicating no or minimal depression (PLY = 57.7% vs DNP = 31.3%), whereas the DNP athletes were more likely to report scores of 10 to 27, indicating more moderate to severe depression than the PLY group (DNP = 40.1% vs PLY = 18.2%) as shown in Figure 2. The DNP group reported a higher (worse) GAD-7 score (mean, [95%CI]) than the PLY group (8.4 [7.2, 9.5] vs. 3.2 [2.2, 4.3], < 0.001) as well as a higher (worse) PHQ-9 score than the PLY group (7.6 [6.4, 8.8] vs. 3.9 [2.8, 4.9], < 0.001). The total GAD-7 and PHQ-9 scores for both groups are found in Table 2.

**Table 2.** Comparison of Anxiety, Depression, Physical Activity and Quality of Life Scores for Adolescent Athletes Who Did or Did Not Play a High School Sport in the Fall 2020

| Variable | All Participants<br>(N = 559) | Did Not Play a Fall<br>Sport (n=388) | Did Play a Fall<br>Sport<br>(n=171) | P-value |
| --- | --- | --- | --- | --- |
| **GAD-7 Total Score** | 6.5 (5.8, 7.2) | 8.4 (7.2, 9.5) | 3.2 (2.2, 4.3) | < 0.001 |
| **PHQ-9 Total Score** | 6.3 (5.6, 7.1) | 7.6 (6.4, 8.8) | 3.9 (2.8, 4.9) | < 0.001 |
| **PFABS Total Score** | 18.7 (17.9, 19.6) | 16.4 (15.0, 17.8) | 23.2 (22.0, 24.5) | < 0.001 |
| **PedsQL Score** |  |  |  |  |
| Physical Summary Score | 88.4 (86.9, 89.9) | 86.5 (84.1, 88.9) | 92.3 (90.1, 94.4) | 0.004 |
| Psychosocial Summary Score | 79.5 (77.5, 81.6) | 75.9 (72.5, 79.3) | 86.4 (83.3, 89.4) | < 0.001 |
| Total Score | 82.6 (80.9, 84.3) | 79.6 (76.8, 82.4) | 88.4 (85.9, 90.9) | < 0.001 |

Reported as mean (SD), N (%), or estimated mean (95% CI) when controlling for Age, Sex, Teaching delivery (in person, online or hybrid) method, and school % eligible for free or reduced lunch percentage.

**Figure 1.**
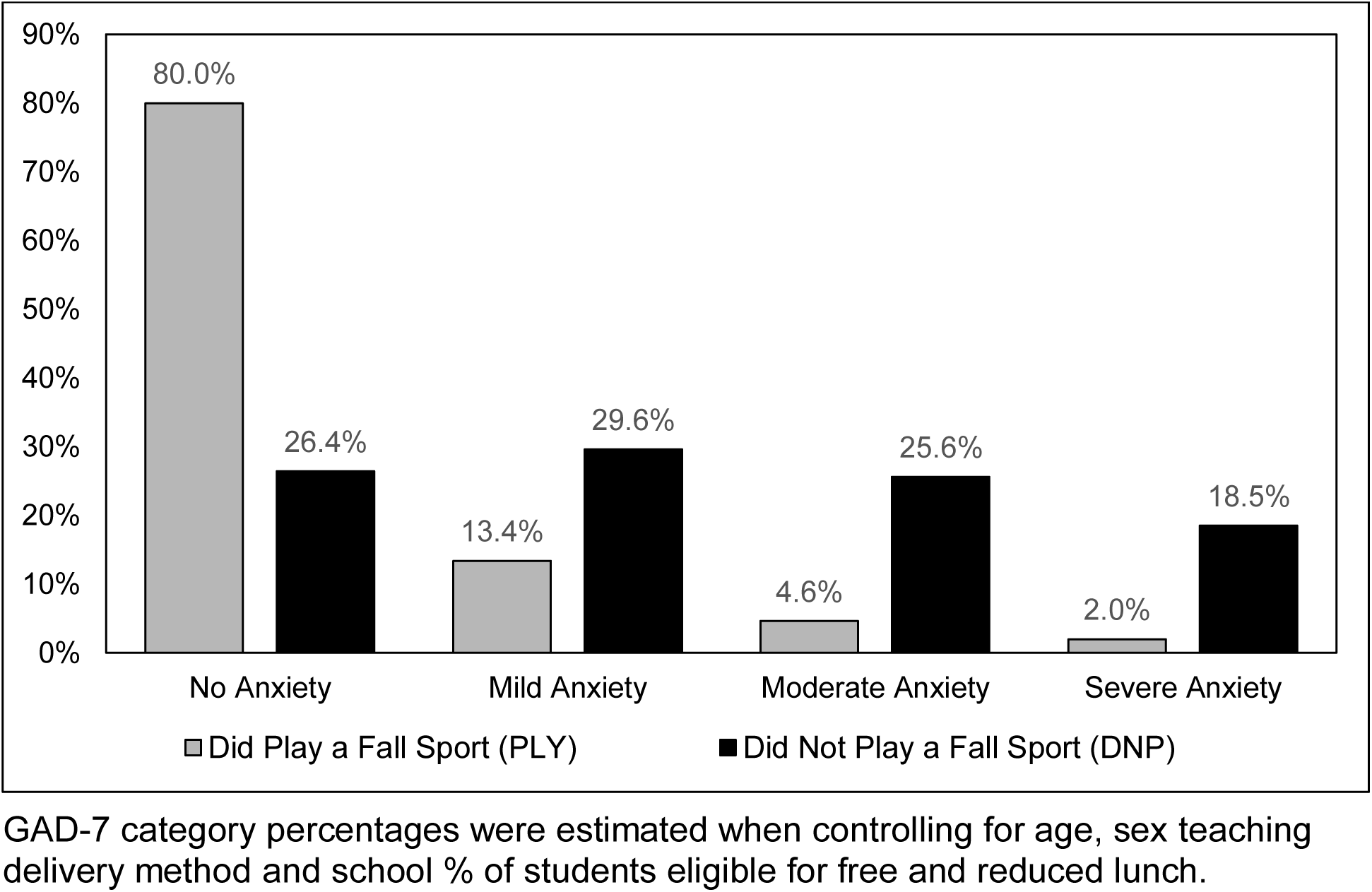
Prevalence of Anxiety Symptoms for Adolescent Athletes Who Did or Did Not Play a High School Sport in the Fall 2020

**Figure 2.**
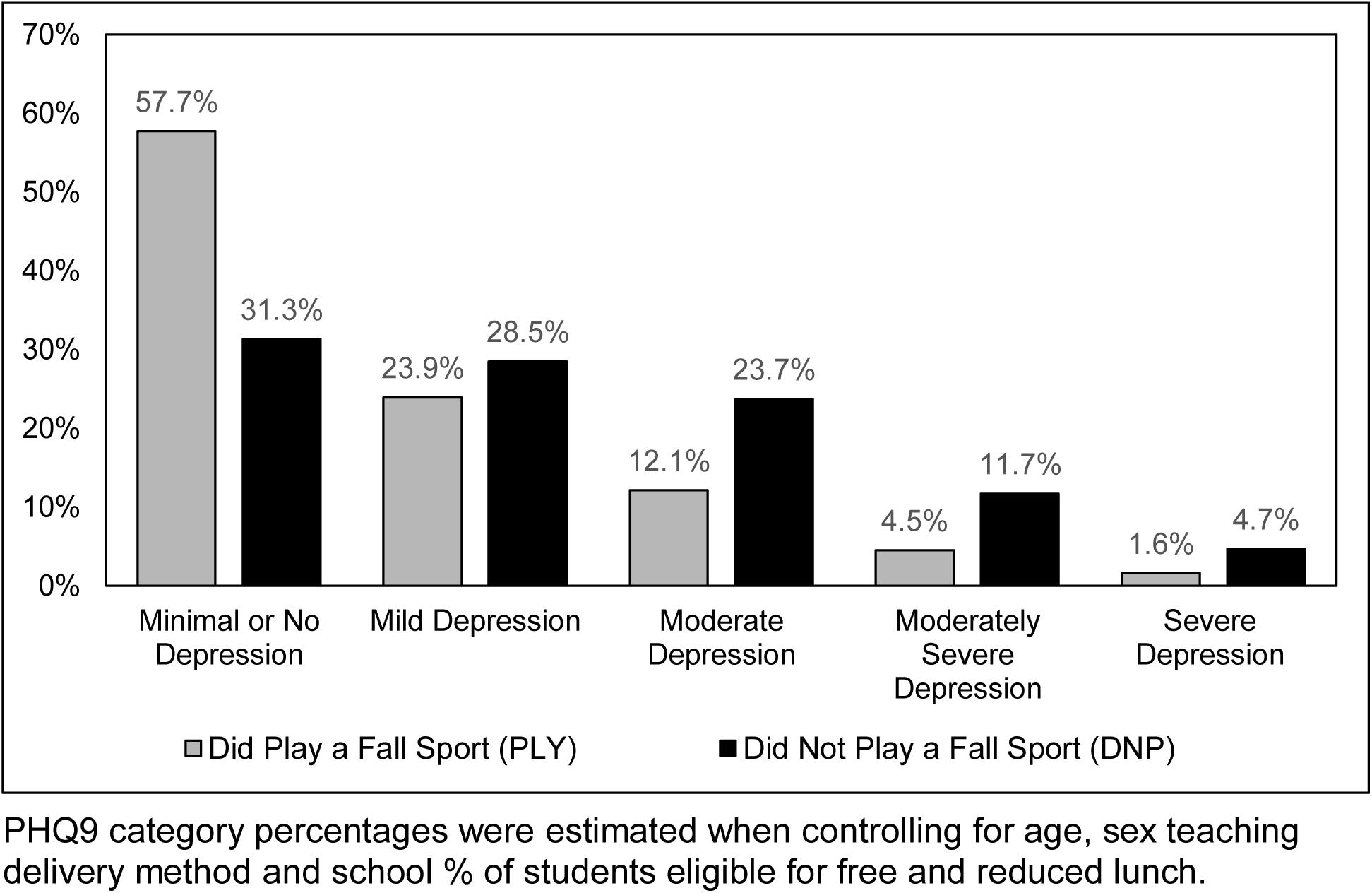
Prevalence of Depression Symptoms for Adolescent Athletes Who Did or Did Not Play a High School Sport in the Fall 2020

### Physical Activity and HRQoL

Physical activity, as measured by PFABS scores (mean, [95%CI]) for the PLY group were 41% higher (better) than the DNP group (PLY = 23.2 [22.0, 24.5] vs. DNP = 16.4 [15.0, 17.8], p < 0.001). The PFABS scores for both groups are found in Table 2. The HRQoL for athletes in the PLY group was higher (better) than the DNP athletes. Specifically, the PedsQL physical health summary scores for the PLY group were higher than the DNP group (PLY = 92.3 [90.1, 94.4] vs. DNP = 86.5 [84.1, 88.9], p = 0.004), as was the psychosocial health summary score (PLY = 86.4 [83.3, 89.4] vs. DNP = 75.9 [72.5, 79.3], p < 0.001) and the total PedsQL score (PLY = 88.4 [85.9, 90.9] vs. DNP = 79.6 [76.8, 82.4], p < 0.001). The PedsQL scores for both the PLY and DNP groups are found in Table 2.

## DISCUSSION

To our knowledge, this is the first study to compare the mental health status, physical activity level and HRQoL between high school athletes who were or were not able to participate in interscholastic sports during the CoVID-19 pandemic. This study builds on prior research demonstrating the dramatic changes in physical and mental health following the cancellation of high school sports in the spring of 2020. ^24,25^ A limitation of the prior studies is the difficulty discerning if the health changes reported were primarily due to the restrictions on sport participation or the result of other factors such as sex, age, socioeconomic status or the lack of in-person school attendance. After controlling for grade, sex, school instructional delivery method and the % of students qualifying for free and reduced lunch, our findings demonstrate that athletes who did not play interscholastic sports experienced significantly worse symptoms of anxiety and depression, lower levels of physical activity and worse HRQoL compared to athletes who did not play sports in fall 2020. This suggests that the re-initiation of sport participation may result in significant improvements in mental and physical health for adolescents during the CoVID-19 pandemic.

### Mental Health

Athletes that played high school sports in the fall of 2020 demonstrated significantly lower symptoms of anxiety and depression than those athletes who did not play a sport. Specifically, athletes in the DNP group were more than 6 times as likely to report moderate to severe symptoms of anxiety and more than twice as likely to report moderate to severe symptoms of depression even after adjusting for age, sex, type of school instruction and % of the students qualifying for free and reduced lunch. This seems to suggest that while PLY athletes continue to demonstrate slightly higher levels of depression and anxiety than historical values, the mental health burden among DNP athletes is considerably worse.^25^ In fact, the DNP group demonstrates levels of moderate to severe anxiety and depression symptoms similar to those identified in a nationwide sample for adolescent athletes in May 2020 (anxiety = 36.7%, depression = 27.1%).^25^ Given the adolescent mental health crisis that existed prior to the onset of CoVID-19, and the known mental health benefits of sport participation, this data seems to suggest that adolescent athletes who are unable to return to sports may be at a significantly greater risk for mental health issues.

Experts have pointed out that the CoVID-19 pandemic has negatively impacted the mental health of youth and may be related to decreased socialization, increased family strain, and reduced access to support services.^13,44^ As such, we recognize that factors beyond sport participation such as the ability to attend school in-person may contribute to adolescent mental health. In addition, depression and anxiety in adolescent athletes have been shown to be related to sex and grade in school. Nonetheless, after controlling for the type of school instructional delivery (online, in-person or hybrid), age and sex, sport participation remained significantly associated with large improvements in anxiety and depression symptoms. Therefore, we can reasonably assume that the increased symptoms we identified among the DNP participants are at least partly attributable to the lack of sport participation.

Our results also support previous research that has demonstrated that sport participation improves the mental health of youth and adolescents.^45-48^ A recent study during the early stages of the CoVID-19 pandemic found that 11.4% of student athletes who were unable to participate in sports reported moderate to severe levels of depression symptoms, which was four times higher than the rate (2.8%) reported by student athletes prior to the CoVID-19 pandemic.^25^ This supports the premise that sport participation may represent an important mechanism to improve the mental health of adolescents as society continues to attempt to mitigate the impact of CoVID-19 in the months and years to come.

### Physical Activity

Our study demonstrates that during the CoVID-19 pandemic, high school athletes who played high school sports had a significantly higher level of physical activity than athletes who did not play a sport. Notably, the total PFABS score for the PLY group was similar to scores (mean, [95%CI]) for healthy high school aged athletes prior to CoVID-19 (24.7 [24.5,24.9]).^25^ Similarly, the PFABS scores for the PLY group were similar to scores for adolescents reported by Donovan (23.8 ± 5.3) as well as to normative data reported by Fabricant (20.2 ± 7.2).^40,49^ Further, the PFABS scores for the PLY group were nearly twice as high as those reported by high school athletes unable to play any sports in May 2020 (12.1 [11.7, 12.5]). Finally, the DNP group reported scores that were 25% and 45% lower than scores reported by Fabricant and Donovan, respectively.^40,49^. This may indicate that by being able to play a sport, these athletes mitigated, to some degree, the low level of physical activity reported during the initial onset of the CoVID-19 pandemic.

Physical activity is known to have a beneficial effect on a wide range of health outcomes in adolescents, including sleep, academic success, well-being, and mental health.^5,6,9,12,45-48,50^ Therefore, it is possible that the identified decrease in mental health in the DNP group may be at least partly due to the removal of the positive effects that physical activity has for adolescents. In addition, childhood obesity was a public health crisis before CoVID-19, and is projected to become worse due to the pandemic.^19,21,51^ Decreased physical activity in adolescents may also have long-term negative effects and implications in terms of increased risk for obesity and cardiometabolic disease if these levels remain low for prolonged periods.^52^ Chronically low levels of physical activity may also compound the mental health consequences of the current crisis and increase the risk ^46,50^ Returning high school sport opportunities is a complex issue and requires careful consideration. Stakeholders should consider the promotion of physical activity for adolescents a top priority during the CoVID-19 pandemic.

### Health Related Quality of Life

HRQoL is a measurement of well-being, and well-being has been associated with self-perceived health, longevity, and healthy behaviors.^53^ We found that adolescent athletes who had returned to sport in fall 2020 reported significantly higher HRQoL than those athletes unable to return to sport. This is consistent with prior studies that have shown that individuals with increased physical activity and/or interscholastic sport participation report higher HRQoL scores compared to inactive adolescents and high school non-athletes.^54-57^ Therefore, it is not surprising that HRQoL scores for the PLY athletes were higher than the DNP athletes.

The total PedsQL scores reported here for the PLY athletes are significantly higher than scores (mean [95%CI]) reported by high school athletes when sports were cancelled (76.7, [76,0, 77.5]). ^24^ Further, the total PedsQL scores reported by the PLY athletes are similar to scores (mean *±* SD) recorded in 14 to 18 year-old athletes prior to the pandemic by Lam (89.4 ± 9.6 to 90.5 ± 10.2), Snyder (89.5 ± 10.1 to 93.6 ± 7.6) and McGuine (90.9 [90.5,91.3] ^25,55,56^ The increased scores for the PLY group may also be attributed to aspects of sport participation beyond the opportunity for structured physical activity such as goal setting, emotional support and social interaction with teammates. This may suggest hat by playing a sport, adolescents may be able to return to a “normal” or “expected” level of HRQoL similar to those recorded prior to the CoVID-19 pandemic.

On the other hand, the total PedsQL scores for the DNP athletes were lower than reported Peds QL scores in non-athletes (84.7 ± 12.7 to 85.8 ± 11.4) reported by Lam et. al.^56^ Interestingly, the DNP scores are similar to scores reported in populations of youth age 5 – 18 with chronic conditions such as asthma (74.8 ± 16.5), cardiac disease (77.4 ± 14.5) and diabetes (80.3 + 12.9) reported by Varni.^58^ The reduced HRQoL in the DNP group may be due in part to the decreased physical activity levels we noted previously. Since HRQoL is multifactorial, however, the scores reported by the DNP group are also likely impacted by other aspects of the lack of sport participation such as the continued loss of social interaction and loss of identity. Nonetheless, this difference persisted after controlling for school instructional delivery type, grade, sex, as well as the % of students eligible for free and reduced lunch, suggesting that the differences we noted may be attributable to the lack of sport participation itself. Further, the fact that the PLY group reported HRQoL similar to data collected prior to the pandemic may indicate that providing sport opportunities can positively impact multiple dimensions of adolescent health. Moving forward, validated health and well-being measurements provide a common metric for future studies to compare their results with our reported data.^53^ These metrics may help policy makers with future decisions that may affect the health and well-being of student athletes in the coming months and years as we transition beyond the immediate impacts of the CoVID-19 pandemic.^59-61^

### Limitations

This study has several limitations. First, the data provided were self-reported from online surveys and not the result of a clinical examination conducted by a health care provider. However, our findings are consistent with reports from experts and US Centers for Disease Control who have stated that CoVID-19 will impact the mental and physical health of youth.^13,14,62^ Second, we acknowledge that there may be a response bias for participants. We cannot know for certain if the sample is representative of all the athletes at the schools or biased towards athletes who were more likely to respond if they experienced the most profound impacts on their health. Third, due to the survey delivery method, our sample may be biased towards athletes from higher socioeconomic groups with easy access to internet services.

Fourth, we used the % of students eligible for free and reduced lunch for each school as a measure of economic status for individual respondents. However, we felt that this would be more accurate as a measure of economic status rather than asking students to accurately report their household income. Finally, we recognize that mental health and overall well-being are complex and are potentially affected by other factors for which we were not able to account in our study and which could confound the results. Specifically, we did not ask the respondents whether they had any fears of contracting the CoVID-19 virus. Nonetheless, we were able to control for sex, age, and school instruction and the % of kids eligible for free or reduced lunch to help better define the impacts of sport participation during CoVID-19.

## CONCLUSION

In this study, we found that adolescent athletes who were able to return to sport participation in fall 2020 reported dramatically lower symptoms of anxiety and depression, higher physical activity levels and higher HRQoL, even after adjusting for grade, sex, and the type of school instructional delivery. As we continue through the CoVID-19 pandemic, it is our hope that this information will help inform public health experts, school administrators, and sports medicine and mental health providers regarding the potential physical and mental health benefits associated with the participation in organized sports for adolescents. We recognize that returning to organized sports is a complex issue and requires careful consideration. Nonetheless, research continues to suggest that sport participation during the CoVID-19 pandemic is associated with significant mental and physical health benefits in adolescents.

## Data Availability

The data is available upon request from the study team.

## Acknowledgements

We gratefully acknowledge and thank all of the student athletes who participated in this study.

